# Exploring Healthcare Professionals’ Perspectives on Artificial Intelligence in Palliative Care: A Qualitative Study

**DOI:** 10.1101/2025.06.22.25330072

**Authors:** Osamah Ahmad, Stephen Mason, Sarah Stanley, Amara Callistus Nwosu

## Abstract

**Background:** The use of Artificial Intelligence (AI) methods in palliative care research is increasing. Most AI palliative care research involves the use of routinely collected data from electronic healthcare records patient data; however, there is little data about the of the views of palliative care healthcare professionals on the role of AI in practice. Determining the opinions of palliative care healthcare professionals on potential uses of AI in palliative care will be useful for policymakers and practitioners to determine, and inform, meaningful use of AI in palliative care practice.

**Aim:** To explore palliative care healthcare professionals’ views on the use of AI for analysis of patient data in palliative care.

**Methods:** Phenomenological study using qualitative semi-structured interviews of palliative care healthcare professionals with a minimum of one-year clinical experience in a hospice in the Northwest of England. Data were analysed using inductive thematic analysis.

**Results:** We interviewed six palliative care professionals comprising of doctors, nurses and occupational therapies. AI was viewed positively, although most participants had not used this in practice. No participants had received training in AI and said that education in AI would be beneficial. Participants described potential benefits of palliative care AI, which included identification of people requiring palliative care interventions, and to evaluate patient experience. Participants highlighted security and ethical concerns of AI, related to data governance, efficacy, patient confidentiality and consent issues.

**Conclusions:** This study highlights the importance of staff perceptions of AI in palliative care. Our findings support the role of AI in enhancing care, addressing education needs, and tackling trust, ethics, and governance issues. This study lays the groundwork for guidelines on AI implementation, urging further research on methodological, ethical, and practical aspects of AI in palliative care.

**Key message:** *What is already known about this topic?:*

- The use of Artificial Intelligence (AI) methods in palliative care research is increasing.
- Most AI palliative care research involves the use of routinely collected data from electronic healthcare records patient data
- There is little data about the of the views of palliative care healthcare professionals on the role of AI in practice.

*What this paper adds:*

- This study provides an overview of the views of specialist palliative healthcare professionals about AI in palliative care, which adds knowledge to a limited evidence base.
- Concerning the use of AI in palliative care, it is important to consider the opportunities of its use, the education needs of staff, the importance of human connection, and practical issues relating to trust and ethics.

*Implications for practice, theory or policy:*

- Future research on palliative care AI should establish standardised reporting for AI studies, seek external validation, and consider ethical issues, which are needed to ensure palliative care AI is safe, fair and reliable.
- Future research should explore the views (about palliative care AI) from different perspectives, including the multidisciplinary team, management, patients, caregivers and other relevant stakeholders.

## Background

Artificial Intelligence (AI) is the science and engineering of creating ‘intelligent’ machines (computers) through developed algorithms that replicate the ability of a machine to think and act like a human.[1] AI involves different methodologies (e.g. machine learning, neural networks, deep learning, and natural language processing), which are processes that enable a machine to be trained to act autonomously, with or without human instruction.[2] AI is already demonstrating significant impact in practice, by facilitating interpretation and analysis of large data fields contained within electronic healthcare datasets.[3, 4, 5] Many applications of healthcare AI help to facilitate early diagnosis and management of disease, supporting individuals to maintain their independence.[6] AI is increasingly used in palliative care (a discipline which provides holistic, person-centered support for older people who are living with life limiting illness[7]), with AI-driven approaches used for several purposes; for example, identification of palliative care need,[8] documentation,[9] case-note analysis,[10] symptom assessment[11] and prognostication.[12] Despite the increased focus on palliative care AI, most current AI applications are for non-palliative care populations. Therefore, there is limited evidence to inform the use of AI in routine palliative care practice, as the needs of palliative care patients are more complex when compared to general (medical and surgical) populations.[13]

Furthermore, there is little known about the views of specialist palliative care professionals’ views on the role of AI in palliative care practice.[14, 15] Understanding the views of palliative care practitioners on AI is important to better understand the opportunities, challenges and implementation issues associated with its use.[16] Consequently, this research study aims to explore the views of palliative care professionals about use of AI in practice, to improve policy and practice in using the AI in palliative care practice.[17]

## Aim

To explore palliative care professionals’ views on the use of AI for analysis of patient data in palliative care.

### Method

This study involved Inductive thematic analysis, which is a method of analysing qualitative data where themes emerge directly from the data, without pre-defined codes or expectations. Inductive thematic analysis was chosen as this provided the researcher with flexibility to explore participants perspectives of AI in palliative care, due to the limited research available on this topic.[18] The lead researcher (OA) was a final year medical student who was supervised by SM and ACN. The study is reported adhering to the COREQ (COnsolidated criteria for REporting Qualitative research) Checklist.[19]

### Study setting

The study took place in a hospice in the North-West of England, a specialised healthcare facility providing care for individuals in the advanced stages of a terminal illness or approaching the end of their lives. The hospice provides a variety of services including a 15 bedded inpatient unit, day services, outpatient clinics, community outreach, patient and family support and bereavement services.

### Sampling and recruitment

Recruitment took place between April and May 2022. The study was introduced at the weekly hospice education meeting, giving potential participants the opportunity to ask questions and express interest in taking part. Study adverts were placed around the hospice, and an email was sent to all hospice staff outlining the inclusion and exclusion criteria for the study. Inclusion criteria were as follows: Palliative care healthcare professionals, working in the hospice, with a minimum of one-year clinical experience during the study data collection period of April – May 2022.

### Data collection

Over a period of 2 months semi-structured interviews were carried out with palliative care healthcare professionals. Participants received written information prior to the interview and provided written consent. Interviews were held face-to-face in a meeting room at the hospice. All interviews were audio recorded using a digital voice recorder. Interviews lasted between 35 and 60 min, and participants were informed that they could pause or discontinue the interview at any time. Each interview began with the researcher providing the participant with background information about the study, and a definition of both artificial intelligence in palliative care. The researcher conducted interviews using an ‘interview guide’, which was used to encourage structure across interviews but also facilitated flexibility, by allowing participants to talk freely about their experiences (Appendix 1). Open questions were used, and the interview schedule was adapted throughout the course of data collection to reflect emergent themes and concepts (Table 1). Field notes and reflections were written throughout the interview process to help make sense of data during the analysis phase.

**Table 1.**
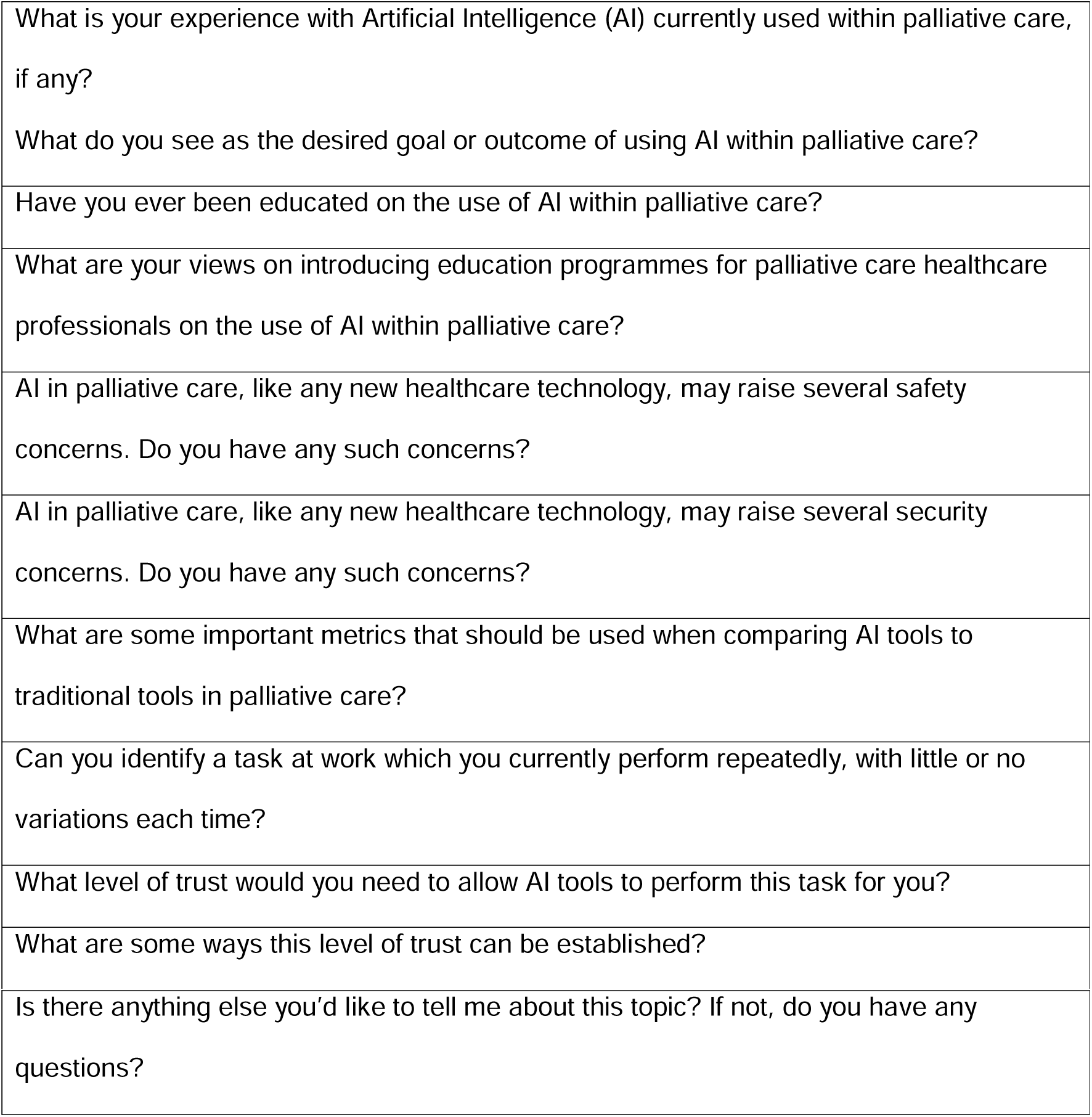
Examples of the interview questions.

**Table 2:**
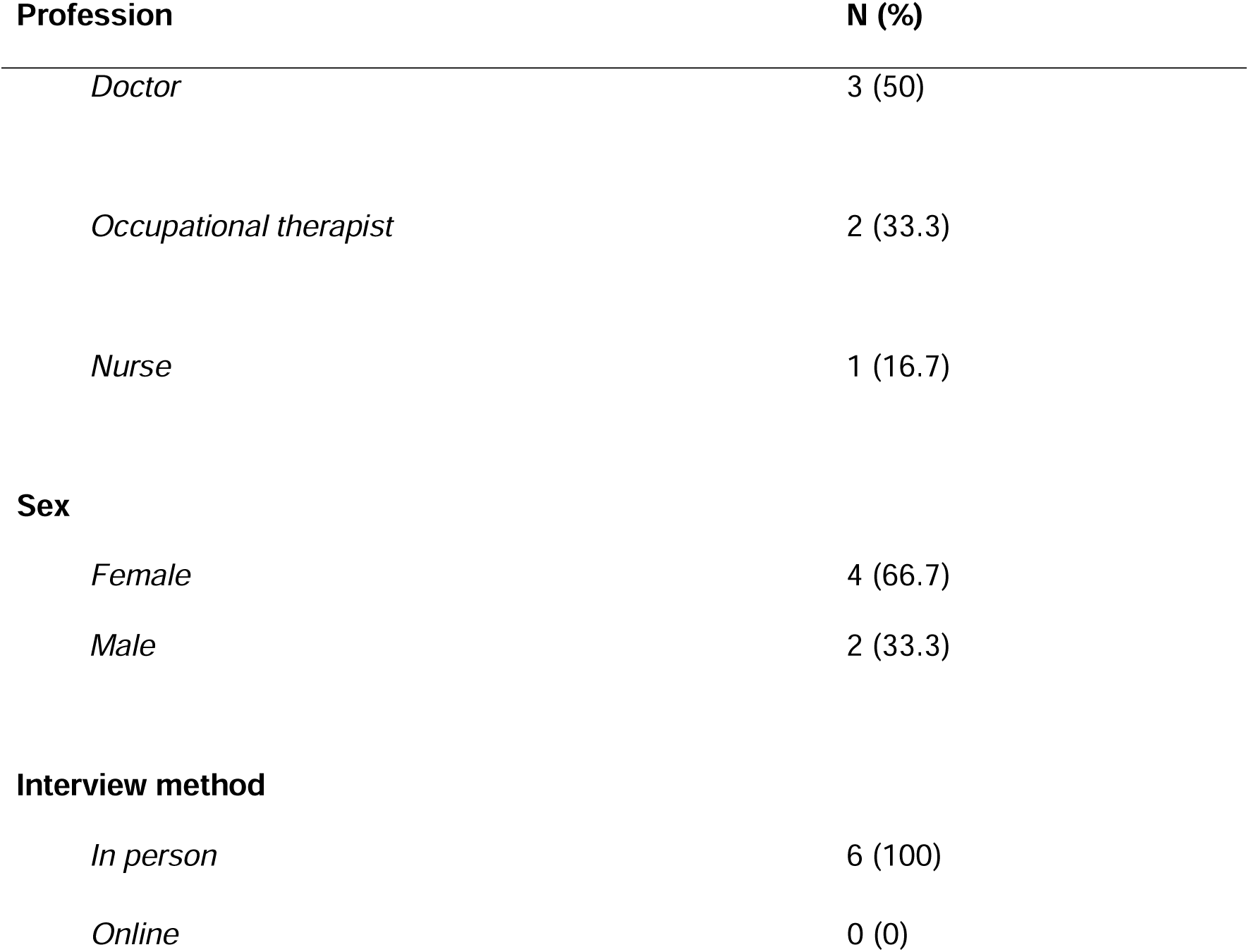
Characteristics of participants.

Recorded interviews were transcribed verbatim by OA. Data were exported to Microsoft word and manual thematic analysis coding was used to systematically label qualitative data extracts to identify patterns and themes. OA used a 6-step thematic analysis proposed by Braun and Clarke,[20] which involves (1) familiarisation with the data, (2) generation of initial codes, (3) development of initial themes, (4) reviewing themes, (5) defining and naming themes and (6) writing up the analysis. Inductive analysis was used with line-by-line coding of participants’ interviews allowing codes to be generated from the data and organised into themes (Appendix 2). Coding was carried out by OA, and regular meetings with ACN were conducted during data collection and analysis to discuss initial findings, evaluate data, challenge emerging ideas and ensure that OA maintained reflexivity throughout the analysis process. Participants were given the opportunity to discuss the developing themes. A consensus was reached to stop data collection on completion of six interviews as the theoretical categories became saturated, and new data was not providing insight or new properties to the categories.[21, 22]

## Results

We interviewed six palliative healthcare professionals, which consisted of one (16.7%) nurse, two (33.3%) occupational therapists and three (50%) doctors. Four (66.7%) participants were female, two (33.3%) were male. All interviews were done in person, face-to-face.

Three themes were developed from the data. These themes were: (1) Opportunities for practice and the need for education, (2) enhancing human care and connection (3), trust and ethical considerations (Figure 1).

**Figure 1:**
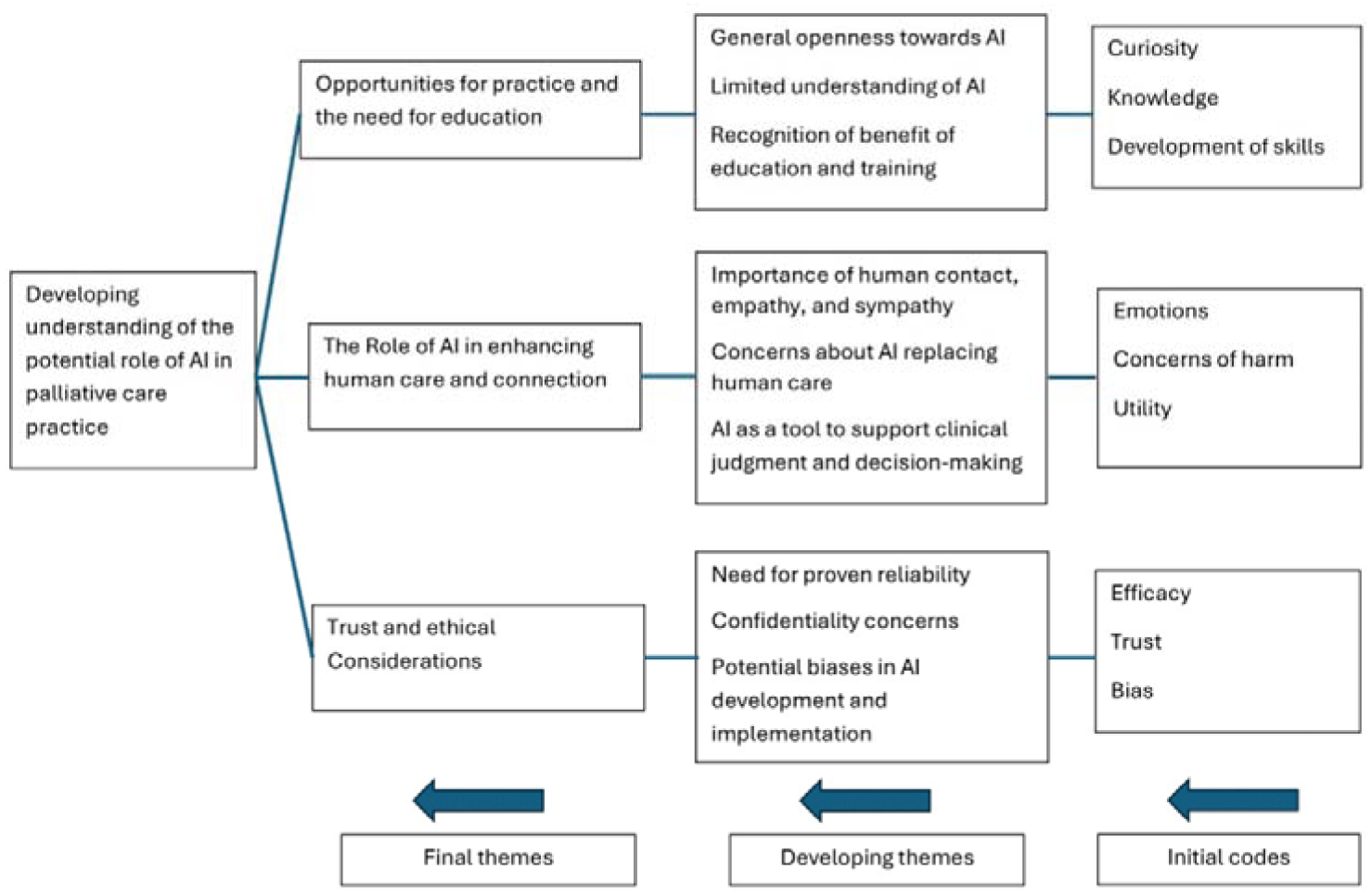
Thematic map showing three main themes

### Opportunities for practice and the need for education

Participants spoke about their openness to consider using AI in palliative care practice, if its use could help to improve care delivery, and efficiency, for patients. Participants spoke of how AI is increasingly being used in other clinical specialities and how, similarly, AI can be used to improve palliative care.

> *“I mean, I think obviously the future is, is going that way, isn’t it? You know, artificial intelligence is being developed at all areas.” (Participant 3)*

Participants spoke about the lack of AI use in palliative care compared to other medical and surgical specialities.

> *“I suppose it’s kind of an alien concept, isn’t it? I think in terms of technology that we use day-to-day at the moment, we’re kind of behind the curve, I would say in, in the NHS, and certainly in this environment. So, I think it’s difficult to kind of visualise what that would be like in kind of day-to-day practice.” (Participant 6)*

Participants described how AI could potentially be used to improve palliative care, describing how AI analysis of electronic healthcare records could help to better identify people with palliative care needs, automate documentation, facilitate data analysis, and provide personalised recommendations for care. Participants provided the following examples when asked how they imagine AI could be used in palliative care practice.

> *“I guess, you know, looking through the hospital records in the hospital. When I see a new patient, I’m manually looking back through the notes to try to find if they’ve had previous encounters with the palliative care team, how times they’ve been in hospital, looking for all the medications to work out what medications they’re on and if any have been stopped.” (Participant 3)*

> *“A good thing would be if something could analyse the entire database every day and say,”look, these are the patients who are flagging up, particularly symptomatic“, or”are using lots of PRN (as required) medicines“or, you know, certain keywords trigger urgent review?” (Participant 4)*

Participants said that they had limited understanding of the different types of AI-driven analytics tools and interventions, which could be used in clinical care. Therefore, participants spoke about the benefits of healthcare professionals receiving education about AI in palliative care.

> *“I think for healthcare professionals, like for me, who’ve maybe not come across so much, or don’t understand it so much, it’s more about knowing about it, and understanding how we can help use it a bit better.” (Participant 1)*

Participants spoke about the importance of education and training to learn about AI in clinical care.

> *“I think that’s only a positive thing. And, I think, to introduce education to help people understand even the basics of what AI is is really important, because I think until we know more about what it is and understand more about what it is, we can’t think of the ways in which we can improve care for the patient.” (Participant 1)*

### Enhancing human care and connection

Participants emphasised the importance of human contact, empathy, and sympathy in palliative care, expressing concerns about AI potentially replacing human care. This is illustrated by the comments of one participant.

> *“I think in palliative care, you know, our strength is based on human connections now think. I don’t mean to be like negative towards, kind of, AI technology and stuff, but I think, actually, you know, the majority my day, when I’m speaking to patients, it’s about human connection.” (Participant 6)*

Participants identified how AI systems could be potentially designed and modelled, to dynamically learn from healthcare professionals and service users, with the objective to develop algorithms which are meaningful in helping palliative care healthcare professionals to provide clinical care.

> *“I think it should be modelled as if, you know, the algorithm behind it is trying to learn, and it’s asking you for confirmation. You know, so it’s saying,’this is what I’ve found’, you know, and you then have to decide whether this is useful, or’can you confirm this is correct?’, or something like that. It should be this kind of back and forth.” (Participant 4)*

Participants described the potential for AI-human partnerships, where palliative care healthcare professionals routinely work with AI-driven data to support care and inform clinical decision-making (e.g. Analysis of electronic healthcare record data to identifying patients with palliative care needs, automated transcription of consultations, personalised treatment recommendations). Participants said that AI could be used as a tool to support clinical judgment and decision-making while maintaining a patient-centred approach.

> *“I guess if you use the program alongside, like, have a step-by-step approach, so you could use the program or the intelligence alongside, you know, a human in kind of partnership. And then gradually, you just have to learn that trust and become more used to it.” (Participant 2)*

### Trust and ethical considerations

Participants said it was important that healthcare professionals are confident that AI-driven clinical tools are trustworthy and reliable. Participants framed their opinions of the trustworthiness of AI in the context of their clinical responsibility of providing care for people with serious illness (who are often consider vulnerable). Participants described the importance of research, to generate the evidence based necessary to ensure that could be safely used in palliative care contexts.

> *“It’d be track record and experience, wouldn’t it? So, you probably would want to have enough evidence to show that it made good decisions.” (Participant 5)*

Participants said that confidentiality was important, with participants expressing concerns related to data privacy and security of patient information, due to integration of AI-driven tools in clinical practice. They emphasised the importance of ensuring that patients’ data remains secure and protected when using AI in palliative care.

> *“I suppose, privacy and data is the main concern as quite often it is in health care, isn’t it? And you know, making sure that data is secure and, you know, things like hacking aren’t an issue and people can’t access patients’ and relatives’ private data easily.” (Participant 1)*

> *“I think with any new technology to have to make sure it’s secure…And, you know, privacy concerns and breaches of confidentiality-So I think all those things are really important.” (Participant 6)*

Participants talked about the risk of for bias associated with AI analysis, which could create (and widen existing) inequalities in palliative care. Participants discussed the importance of addressing biases in AI algorithms and considering public perception when considering how AI can be best used to support palliative care. Participants said that AI-driven metrics should be used to benefit palliative care patients, and its use should be used inclusive, unbiased, and ethical.

> *“If our algorithms have been driven by developers in Silicon Valley in California and most of them are white, male and young, and the modelling has been tested on a particular set of individuals, which don’t have certain characteristics, which mean that certain people are not represented, you then might get a device or an algorithm which isn’t tailored for the needs of certain people.” (Participant 4)*

### General comments of participants

Overall, AI was viewed positively by many participants, although many said they had not used it in practice. No participants had received training in AI, and all participants commented that relevant formal education in AI would be beneficial. Participants described potential benefits of palliative care AI, which included identification of people requiring palliative care interventions, and to evaluate patient experience. Participants highlighted several security and ethical concerns related to data governance, patient confidentiality, psychological harm to patients, accuracy of AI decisions and consent.

## Discussion

Concerning the use of AI in palliative care, it is important to consider the opportunities of its use, the education needs of staff, the importance of human connection, and practical issues relating to trust and ethics.

### Importance and uniqueness of this paper

This study provides an overview of the views of specialist palliative healthcare professionals about AI in palliative care, which adds knowledge to a limited evidence base. Our qualitative approach facilitated in-depth exploration of participants experiences, which captured the participant’s nuances and complexities. The evidence derived from this study improves knowledge about the views palliative care staff have about AI, which will shape its clinical use. Evidence demonstrates that improved staff involvement can improve effectiveness, and success, of healthcare systems interventions.[23, 24]

### Relation to previous work in this area

In this study, staff positively described potential opportunities that AI could be used to support palliative care, with themes consistent with previous work conducted with generalist staff.[25] In our study, staff identified several hypothetical possibilities where AI could be positively used to improve their practice (e.g. predictive modelling, text screening, symptom assessment, communication etc etc); which are consistent with current AI developments of AI in palliative care.[26] Our findings align with previous research, that recommends that healthcare professionals receive formal education and training in AI.[14, 27] Specifically, previous research advises for palliative care education programmes to include holistic overviews of AI technologies, ethical considerations, and case studies that highlight real-world applications and challenges of AI in palliative care.[28]

Our findings support previous work, which describes the importance of focusing on human connection in palliative care, and ensuring that AI tools are meaningfully used to improve the experience of patients, caregivers and staff.[25] Consistent with previous work, we highlight potential problems, and bias, which may occur from using AI in palliative care, due to limited evidence of patient-centred outcomes measures in palliative care populations.[29] In our analysis, participants described the ethical challenges of using AI in palliative care, which are similar to the findings of previous work.[28] These ethical challenges included themes of promoting transparency and accountability in AI systems, regular ethical review and continuous impact assessments, ensuring patient autonomy and informed consent and safeguarding data privacy and security.[28] Consequently, our data supports previous studies reporting the importance of making complex AI models more understandable and trustworthy to enable non-expert users to see how the AI models process input and produces outputs.[30]

### Limitations

This study is small, focused on one hospice in the North West of England, meaning that the findings may not be generalisable to other palliative care settings (e.g., home, the community, hospital and nursing homes). This study lacks representation of some professional roles (e.g., social work, spiritual care, pharmacy and fundraising), which means we lack data about how AI may impact wider specialist roles in the palliative care multidisciplinary team. Furthermore, our study did not include the perspectives of patients, caregivers, and other relevant stakeholders.

### Importance to policy, practice and research

Decision makers should consider the perspectives of palliative care staff, when developing, and implementing, AI tools for use in palliative care. When exploring the role of AI in palliative care, decision makers should consider opportunities to improve care, focus of human needs and address ethical and governance issues. For education, is important that palliative care professionals receive education about AI and its integration in practice. This may include the development of formal undergraduate and postgraduate training curricula, which include modules on AI and associated topics, such as ethics and governance.[17] [28]

Future research on palliative care AI should establish standardised reporting for AI studies, seek external validation, and consider ethical issues, which are needed to ensure palliative care AI is safe, fair and reliable.[30] Future research should explore the views (about palliative care AI) from different perspectives, including the multidisciplinary team, management, patients, caregivers and other relevant stakeholders. Researchers should involve interdisciplinary partnerships and collaboration, to facilitate work across essential interconnected themes such as design, computing, data analysis, ethics and translational medicine.[14] [31, 32]

## Conclusions

This study shows the importance of considering the staff perceptions of the role of AI in palliative care. Our findings support growing evidence about the development, and use, of AI in palliative care practice. Our findings demonstrate the importance to consider opportunities to use AI meaningfully to improve human focused care, the education needs of staff, and the need address practical issues relating to trust, ethics and governance. This study can serve as a foundation for developing guidelines for AI implementation in palliative care, addressing various stakeholders, from healthcare professionals to policymakers. Future research should study methodological, ethical and practical issues to determine how AI technology can best support delivery palliative care to those with serious illness.

## Data Availability

All data produced in the present study are available upon reasonable request to the authors.

## Acknowledgments

We would like to thank the participants for their valued input and participation in this study. We thank the hospice management and research governance group for supporting study recruitment. SS and ACN’s roles are funded by Marie Curie.

## Ethics and consent

The University of Liverpool ethics committee gave ethical approval for this work. This study was approved by the hospice research governance group. All participants provided written informed consent.

## Author Contributions Statement

OA designed the study, conducted the research, wrote the manuscript. SM, SS and ACN designed the study, provided oversight of the project and wrote the manuscript. All authors reviewed and revised the manuscript.

