## Appendix 1 for "Exploring Healthcare Professionals’ Perspectives on Artificial Intelligence in Palliative Care: A Qualitative Study"

**Artificial intelligence in palliative care: a study on the views of palliative care healthcare professionals on the role of artificial intelligence in the analysis of healthcare data for people with palliative care needs.**

**Narrative Stimulus:**

We are interested in the views of Specialist Palliative Care Healthcare Professionals (SPC-HPs) on AI use within palliative care. For this interview, the term AI refers to machines with human-like cognitive abilities, such as thinking, knowing, remembering, judging, and problem-solving.

May I ask what you understand about AI in the context of palliative care? I would like to follow up this question by exploring this in further detail. Please take your time and feel free to say anything and as much as you want regarding the topic.

Current AI research in palliative care is largely focused on mortality and survivability prediction, and on natural language processing of free-text patient notes.

**Interview Guide:**

- What is your experience with AI currently used within palliative care, if any?
- What do you see as the desired goal or outcome of using AI within palliative care?
- Have you ever been educated on the use of AI within palliative care?
- What are your views on introducing education programmes for SPC-HPs on the use of AI within palliative care?
- AI in palliative care, like any new healthcare technology, may raise several safety concerns. Do you have any such concerns?
- AI in palliative care, like any new healthcare technology, may raise several security concerns. Do you have any such concerns?
- What are some important metrics that should be used when comparing AI tools to traditional tools in palliative care?
- May I ask you, if you can, to identify a task at work which you currently perform repeatedly, with little or no variations each time?
  - What level of trust would you need to allow AI tools to perform this task for you?
  - What are some ways this level of trust can be established?

Is there anything else you’d like to tell me about this topic? If not, do you have any questions?
