## Appendix 2 for "Exploring Healthcare Professionals’ Perspectives on Artificial Intelligence in Palliative Care: A Qualitative Study"

### Theme 1: Openness and Education in AI

#### Awareness of own knowledge

1. "…I'm very limited with understanding how it could be used to improve things for people a little bit more. I think that's something that, you know, I definitely need to learn a bit more about."
2. "A couple of years ago, there was an interest there though, a fascination. I do feel very positive about it."
3. "I haven't [received AI education]."

#### Positivity towards AI education

1. "I think it's definitely important that we start getting more education and understanding what it is so that we can be the people who, you know, create ways to use it better for our patients and talk to our patients about how we can use it better for them and how we can improve things for them with the use of AI."
2. "I think if we are starting to look at artificial intelligence in healthcare, I think it is the training side of it."
3. "I mean, I think obviously the future is, is going that way, isn't it? You know, artificial intelligence is being developed at all areas."
4. –
5. –
6. "Yeah, I mean, I think that'd be, that'd be really well, cool."
   a. "No, I think it's good to be educated on topics. Absolutely."

#### Belief in the inevitability of AI integration

1. –
2. –
3. –
4. –
5. –
6. "I think, without a doubt, these technologies are going to have a massive impact."

#### Potential applications of AI in Palliative Care

1. "Using things like the documentation that we would have to do on a patient every day on a daily basis. We've got templates on our electronic record system, so we're filling in the same templates every day, at the roughly the same time a day, to give an update on what care we've given to the patients on that day, what that involved and what we've done for them."
2. "So, an analytic tool will be very good. And I suppose I say that because I find all of that type of analysis of computer-based things quite boring and quite mundane."
3. "I guess, you know, looking through the hospital records in the hospital. When I see a new patient, I'm manually looking back through the notes to try to find if they've had previous encounters with the palliative care team, how times they've been in hospital, looking for all the medications to work out what medications they’re on and if any have been stopped."
4. "A good thing would be if something could analyse the entire database every day and say, “look, these are the patients who are flagging up, particularly symptomatic”, or “are using lots of PRN medicines” or, you know, certain keywords trigger urgent review?"
   a. "Particularly in elderly care, when the use of data is being talked about for what the Americans call ageing in place, and helping to keep people in the home environment, you're looking at data that's being collected about patients in their own homes."
5. "But I guess it's more about making it easier to document or to communicate with other professionals."
   a. "When we’re documenting notes. But I guess that's different every time isn't it? But it's a pattern of- You do the same kind of- You document things in the same way. I guess the content will be different. I say that probably, you know, the task that takes the most time away from patients I would say."
6. "I think people are comfortable, you know, being in that kind of virtual environment and, kind of- I don't know if you know many gamers, but they just-They could quite happily be in that kind of virtual environment for a long period of time and actually might feel more comfortable interacting with people in that way."

### Theme 2: The Role of AI in Enhancing Human Care

#### Importance of human contact

1. "It could present with some really, you know, difficult situations, especially in palliative care when there's a lot of sensitive information held and, you know, we talk very sensitively to the patients and a lot of the topics that we cover are sensitive topics, so I sort of see that as being: if they didn't understand AI, then it could be quite that could become a risky situation, I suppose."
2. "I was a bit sceptical about some of that care being provided by robots just because of, I suppose, the lack of emotion that I think that type of application would have compared to the human approach."
3. –
4. "I think one of the challenges that people have with artificial intelligence is that they imagine a computer or a robot, which is doing the job of the human being for you."
5. "I suppose it's just the impact it will have on like doctor-patient relationships, that there's more and more of a doctor's work being done by an AI?"
   a. "I feel like, if an AI gets used, you're giving patients information... But we are able to recognise that, and we offer that human kind of sympathy and empathy."
6. "I think in palliative care, you know, our strength is based on human connections now, I think."
   a. "I think ideally, you know, face-to-face contact with the opportunity, you know, for touch is important. I think you can really make a connection with someone, you can build rapport, build trust."

#### Working in partnership with AI

1. –
2. "So, if you did have some type of program, artificial intelligence, that would be amazing."
   a. "But I think if you use the program alongside, like, have a step-by-step approach, so you could use the program or the intelligence alongside, you know, a human in kind of partnership."
3. "Obviously, they still have to use their own clinical reasoning and judgement, but the machine will assess the risk for them."
4. "So, if we collect somebody's data, and it goes into electronic healthcare record, I think it's important for services and practitioners just to be aware about how they can extract that data."
   a. "The language should be talking about helping to support decisions. It's not to be a replacement."
   b. "So, the challenge with an algorithm, using AI, is if it ends up replacing human contact and things become driven according to the algorithm rather than driven to what patients need."
5. "You’d still have to have the human decisions in case, you know."
   a. "So, I think often that happens when there's, you know, multiple patients in a pool and you're trying to make sure you give the most needy patients the right treatment. These are quite subjective, I think, and probably we’re a bit biased in certain ways as humans, which sometimes is okay, but I'm sure other times we make decisions that, actually, are maybe not the right decisions."
   c. "I imagine that the computers would make errors sometimes, I don't know."
6. "I think that's key — clinical judgement, isn't it?"
   a. "So, I think any tool that could achieve that, I think, would probably be welcomed, as long as it doesn't replace human contact."

#### Common goal of patient-centred care

1. –
2. –
3. "We’d want to see that there wasn't a dip in quality of life, that patient goals were still being achieved."
4. "I'd like to see us being more ambitious with our approaches to AI and use more data points to try to get more meaningful data to help clinicians."
5. "Yes, so I think with an AI you want it to do a job that we're not doing well enough, you know, as we are at the moment."
6. "I think most patients want to get in and out of the hospital as quickly as they can with the correct treatments and good care, and with a good follow up plan, and confidence in their treatment."

### Theme 3: Trust & Ethical Considerations

#### Proving reliability of AI

1. –
2. "You’d have to have 100% trust in it, with the appreciation that everything fails."
3. "Check it. Trial run it, check it."
4. –
5. "As in, what would make me trust the AI? It’d be track record and experience, wouldn’t it? So, you probably would want to have enough evidence to show that it made good decisions."
   a. "I think if you were surrounded by colleagues that trusted it, then I think for a lot of people that that would certainly make a difference."
   b. "I suppose maybe knowing its error rates? And is that a one in a million event is that a one in one-hundred?"
6. "I guess you'd want that reassurance that it translated to sick patients or to you know- It would have to be on a trial basis."

#### Confidentiality concerns

1. "I suppose privacy and data is the main concern as quite often it is in health care, isn't it? And you know, making sure that data is secure and, you know, things like hacking aren’t an issue and people can't access patients’ and relatives’ private data easily."
2. "Using technology, it just opens that risk greater, doesn't it, to people who are intelligent enough to break down the security walls?"
3. "I suppose, mainly with regards to kind of data breaches or sharing of information."
4. "What I would really like to see is putting the patient first and looking at the needs and requirements of those individual patients."
   a. "So, for me, it's those questions, it's who owns the data? How secure is the data? What are data being used for? What do people think about it? Are there third parties involved that are doing data analysis beyond to the NHS?"
5. "I think, I suppose, protection of people's data, because I guess the way these AIs work is by looking at large amounts of data which, at a human level, you would struggle to analyse, and they can look at a large number."
   a. "I guess the whole idea is that these AI should have less error than human error."
6. "I guess with any new technology to have to make sure it's secure. I guess the difficulty is, we use EMIS here, but you know, hospital systems use different computer systems, don't they? And that ability to communicate between one another presents its own difficulties."

#### Potential biases in AI development and implementation

1. –
2. "I appreciate there would be precautions that you'd need to take and risks, and it's really difficult to answer knowing that there's so many different variables to that."
3. "I think that would be hard, because I think every patient is so individual."
4. "It really depends on the question that you're asking. So, for example, many of the studies looking at prognosis, trying to identify how long people are going to live in a hospital setting, are using different metrics and different variables to try to predict that. Most of those metrics are variables that they'll be able to measure from a data set. So automatically, you've got a bias there, because you've got various things which can't be measured, which aren’t in electronic datasets, that you can't measure, which have been excluded from your algorithm."
   a. "If our algorithms have been driven by developers in Silicon Valley in California and most of them are white, male and young, and the modelling has been tested on a particular set of individuals, which don't have certain characteristics, which mean that certain people are not represented, you then might get a device or an algorithm which..."
5. "I think probably what would influence me as well is what the public perception of it, because if the public really aren’t on board with it, probably, I'm going to be a more anxious about trusting it because of the because, if there's a negative outcome, and the whole public hate AI, then probably you're going to get a lot more complaints about using it."
   a. "It’s difficult, because I don't know how the programming works. I don't know how robust it is. I presume it's better, but I don't know how often errors occur."
